# A novel morphometric signature of brain alterations in type 2 diabetes: patterns of changed cortical gyrification

**DOI:** 10.1101/2021.02.25.21252196

**Authors:** Joana Crisóstomo, João V. Duarte, Carolina Moreno, Leonor Gomes, Miguel Castelo-Branco

## Abstract

Type 2 diabetes is a chronic disease that creates atrophic signatures in the brain, including decreases of total and regional volume of grey matter, white matter, and cortical thickness. However, there is a lack of studies assessing cortical gyrification in type 2 diabetes. Changes in this emerging feature has been associated mainly with genetic legacy, but environmental factors may also play a role. Here, we investigated alterations of the gyrification index and classical morphometric measures in type 2 diabetes, a disease with complex etiology with both underlying genetic and more preponderant environmental factors.

In this cross-sectional study we analyzed brain anatomical magnetic resonance images of 86 participants with type 2 diabetes and 40 healthy control participants, to investigate structural alterations in type 2 diabetes, including whole-brain volumetric measures, local alterations of grey matter volume, cortical thickness and the gyrification index.

We found concordant significant decrements in total and regional grey matter volume, and cortical thickness. Surprisingly, the cortical gyrification index was found to be mainly increased in cortical sensory areas in type 2 diabetes. Moreover, it correlated with features of metabolic control. Our findings challenge the classical neurodevelopmental association of gyrification mostly with genetic determinants. While we found mainly increased gyrification in more genetically constrained sensory areas in type 2 diabetes, our correlation results concurrently suggest an influence of metabolic control in alterations of gyrification in type 2 diabetes. Further studies should address causal influences of genetic and/or environmental factors in patterns of cortical gyrification in type 2 diabetes.

## 1. Introduction

Type 2 diabetes is a highly prevalent chronic metabolic disease that affects many tissues and organs, and the brain is not an exception. Despite the ongoing controversies on the nature of its impact in the brain, changes in structure and function are well documented in diabetes. This disease is commonly associated with cerebrovascular disease, cognitive decline, Alzheimer’s disease and other types of dementias and depression [Bancks et al., 2017; Cheng et al., 2012; Moheet et al., 2015; Verdile et al., 2015]. Studies of structural magnetic resonance imaging (MRI) with volumetric analysis at the whole-brain level and voxel-based morphometry (VBM) have shown brain atrophy, including lower total and regional volume of grey matter (GM) and white mater (WM) in type 2 diabetes patients when compared to non-diabetic controls [Moheet et al., 2015; Moran et al., 2013]. Decrements in cortical thickness were also already demonstrated with surface-based morphometry (SBM) studies in type 2 diabetes patients [Chen et al., 2017; Moran et al., 2019]. Conceptually, the volume of GM and the cortical thickness seem to be influenced by environmental factors related with diabetes progression, such as patients’ glycemic profile, lipidic status, vascular complications and insulin taking [Brundel et al., 2010; Bryan et al., 2014; Chen et al., 2015; Chen et al., 2017; Shi et al., 2019]. Notably, cortical gyrification is an interesting surface feature that has been suggested to be more specifically determined by genetic and neurodevelopmental factors [Ronan and Fletcher, 2015], which to the best of our knowledge was never assessed in type 2 diabetes.

Gyrification refers to the development of the folding surface patterns on the brain [Welker, 1990; Zilles et al., 1988], being highly heritable, evolutionarily conserved, and similar amongst closely related animal species [Alexander-Bloch et al., 2020; Zilles et al., 2013]. Due to gyrification, the cortical surface area and thus the volume of cortical GM can increase dramatically [White et al., 2010]. It is postulated to be dictated by the genetic signature and the period of greatest development of brain gyrification is during intrauterine development [Papini et al., 2020; White et al., 2010]. Genetic control of this process is now starting to be understood both in animal models using genome editing [Johnson et al., 2018] and also in the human brain [Alexander-Bloch et al., 2020]. A quantitative approach to measure local gyrification is known as the gyrification index, which is a ratio between the length of the outer folded surface of the brain, including sulci, and the length of the outer surface excluding sulci [Schaer et al., 2008; Yotter et al., 2011; Zilles et al., 1988]. The development of this quantification concept has allowed to identify altered gyrification in several diseases with genetic or neurodevelopmental basis as well as in complex etiology diseases [Bearden et al., 2009; Casanova et al., 2004; Gaser et al., 2006; Jou et al., 2010; Lebed et al., 2013; Lin et al., 2007; Palaniyappan and Liddle, 2012; Zhang et al., 2010]. Besides alterations in pathologically impaired brains, there is also evidence suggesting that changes in gyrification also occur due to environmental factors [Amunts et al., 1997; Luders et al., 2012], as well as in healthy ageing [Hogstrom et al., 2013; Lamballais et al., 2020]. However, there is no investigation regarding this cortical feature in diabetes.

We hypothesize that it is conceivable that alterations in the brain surface morphology, namely in gyrification, may also occur in diabetes, thus following the trend of documented structural changes and functional impairments in this complex disease.

In this cross-sectional study we aimed at investigating this novel feature of cortical gyrification in type 2 diabetes, as well as more classical structural measures, including whole-brain volumetric tissue volumes - total intracranial volume (TIV), GM, WM, and cerebrospinal fluid (CSF), local GM volume and cortical thickness.

## 2. Materials and Methods

### 2.1 Participants

After signing the informed consent, 190 participants were enrolled in a cross-sectional study, between 2012-2014, and divided in two experimental groups: a type 2 diabetes group (T2D) and a control group (CNT). Type 2 diabetes patients were recruited at the Endocrinology Department of the University of Coimbra Hospital, diagnosed using standard WHO criteria at the moment of recruitment, based on glucose levels, oral glucose tolerance test and glycated hemoglobin (HbA1c) [WHO, 1999; WHO, 2011]. Inclusion criteria for type 2 diabetes group were: (i) age between 40 and 75 years; and (ii) type 2 diabetes for at least 1 year before the enrolment in the study. Control individuals were recruited from the general population of the hospital or university staff and their relatives. Inclusion criteria for CNT group were: (i) age between 40 and 75 years and (ii) type 2 diabetes diagnosis excluded based on levels of HbA1c and fasting glucose. General exclusion criteria were: (i) history of neurological or psychiatric disorders; (ii) substance abuse/dependence; and (iii) contra-indication for MR imaging.

Demographic and clinical data were collected for all participants by research physicians and fasting blood samples were collected by venous puncture by research nurses for posterior biochemical analysis, according to the hospital standard procedures.

The study was approved by the Ethics Commission of the Faculty of Medicine of the University of Coimbra and was conducted in accordance with the declaration of Helsinki. All participants provided written informed consent to participate in the study.

### 2.2 Magnetic resonance imaging data acquisition

Data were collected at the Institute of Nuclear Sciences Applied to Health - University of Coimbra, using a Siemens Magnetom TIM Trio 3 Tesla MRI scanner with a phased array 12-channel birdcage head coil (Siemens, Munich, Germany). For each participant, 3D anatomical T1-weighted MPRAGE (magnetization-prepared rapid gradient echo) images were acquired with the following parameters: repetition time (TR) = 2530 milliseconds; echo time (TE) = 3.42 milliseconds; inversion time (TI) = 1100 milliseconds; flip angle 7°; 176 single-shot interleaved slices with no gap with isotropic voxel size 1 × 1 x 1 mm; field of view (FOV) = 256 mm.

### 2.3 Data analysis

Out of 190 participants recruited, 17 dropped out before MRI scanning. Additionally, 3 participants were excluded after a quality assessment by visual inspection of acquired structural data, because of severe movement artefacts and anatomical anomalies. Then, aiming to do between-groups analysis balanced for age, 86 type 2 diabetes participants and 40 CNT participants were selected, which is well above recommended sizes for neuroimaging studies [Friston, 2012].

Images were processed and analyzed using SPM12 (Wellcome Trust Centre for Neuroimaging, London, UK; http://www.fil.ion.ucl.ac.uk/spm/software/spm12/) and CAT12 toolbox (Structural Brain Mapping Group, Jena University Hospital, Jena, Germany; http://dbm.neuro.uni-jena.de/cat/), as it offers processing and analysis pipelines for both voxel-based morphometry (VBM) as well as surface-based morphometry (SBM) (including cortical thickness and gyrification index). This toolbox has been previously used and validated in morphometric studies in clinical populations, including by our own group [Madeira et al., 2020].

#### 2.3.1 Voxel-based morphometry

After quality assessment of original images, the images were centered in the anterior commissure and oriented in AC-PC (anterior commissure – posterior commissure) plane. Subsequently, images were spatially normalized into Montreal Neurological Institute (MNI) standard space and segmented into three tissue classes - grey matter (GM), white matter (WM) and cerebrospinal fluid (CSF) – using partial volume segmentation with adaptive maximum a posteriori approach. Total intracranial volume (TIV) was also determined for all scans. The extracted modulated normalized GM maps were smoothed using a 12 mm full width at half maximum (FWHM) kernel and used for further analysis. An absolute masking threshold of 0.1 was applied to the VBM data. The Automated Anatomical Labelling and Yale BioImage Suite (v1.3) brain atlases were used to label the regions with differences between groups, according to their MNI coordinates.

#### 2.3.2 Cortical thickness

Cortical thickness was extracted based on the absolute mean curvature approach [Luders et al., 2006]. Extraction of the cortical surface (using CAT12 standard procedure) resulted in the construction of a mesh of the central surface, i.e., the surface between the GM/CSF border and the GM/WM boundary. The central surface as well as cortical thickness are estimated in one step using a projection-based distance measure [Dahnke et al., 2013]. The vertex-wise cortical thickness measures were resampled and smoothed using a 15 mm FWHM gaussian kernel.

#### 2.3.3 Gyrification index

Local (vertex-wise) gyrification index maps were calculated based on the same algorithm for extraction of the cortical surface implemented in CAT12, as given above for cortical thickness analysis [Luders et al., 2006; Yotter et al., 2011]. The local absolute mean curvature of this central surface was then calculated by averaging the mean curvature values from each vertex point within 3 mm from a given point. In a second step, we applied 15 mm FWHM smoothing to the gyrification index maps.

### 2.4 Statistical analysis

The analysis of demographic, clinical data, and total brain volumes (TIV, GM, WM, CSF) was performed with GraphPad Prism 6. Normality of the data was tested using D’Agostino-Pearson test. Parametric and non-parametric tests were used when data were normally distributed or when assumption of normality was not met, respectively. Parametric t-test and non-parametric Mann-Whitney test were used to assess between-groups differences. Parametric Pearson test and non-parametric Spearman test were used to calculate correlations between disease-related parameters (glucose, HbA1c, age at diagnosis and disease duration) and the gyrification index. Chi-square test was used to assess the difference in gender distribution between groups.

The statistical analyses of imaging data were performed in the CAT12/SPM12 statistical module applying t-test to each of the three morphometric measures: GM volume with VBM, and cortical thickness and gyrification index with SBM. Using age and gender as covariates (and for VBM analyses, additionally, TIV), group differences applying thresholds of *p*<0.05 with family-wise error rate (FWE) correction for multiple comparisons were tested. When the correction was too stringent, and not to miss an exploratory interesting effect, the statistical maps were thresholded at voxel level with p<0.001 and then corrected at the cluster level with cluster extent correction.

## 3. Results

### 3.1 Demographic and clinical data

Demographic and clinical data are depicted in Table 1. The mean age of type 2 diabetes group was 60.67 years and of CNT was 57.73 years and no statistically significant difference between groups was found. Gender distribution between groups was also not different. As expected, type 2 diabetes group showed significantly higher levels of fasting glucose and HbA1c, when compared with controls. The mean age at diagnosis of diabetes was 47.73 years and mean disease duration was 12 years. All patients were taking oral antidiabetic drugs, 81.3% of which in combination with insulin (data not shown).

**Table 1:** Demographic and clinical data

| Number |  | T2D<br>86 | CNT<br>40 | Test | *p* value |
| --- | --- | --- | --- | --- | --- |
| Age (years) | | 60.67 ± 7.89 | 57.73 ± 8.06 | $t = 1.94$ | 0.06 |
| Gender (%) | Male | 59.3 | 50 | $\chi^2 = 0.96$ | 0.33 |
|  | Female | 40.7 | 50 |  |  |
| Fasting glucose (mg/dL) | | 166.5 ± 62.56 | 95.05 ± 10.89 | $U = 295.5$ | <0.0001 |
| Glycated Hemoglobin | (mmol/mol) | 79.53 ± 25.72 | 35.13 ± 3.98 | $t = 10.21$ | <0.0001 |
|  | (%) | 9.43 ± 2.37 | 5.36 ± 0.38 |  |  |
| Age at diagnosis (years) |  | 47.73 ± 10.26 | - | - | - |
| Disease duration (years) |  | 12.05 ± 8.88 | - | - | - |

Data are presented as mean ± SD for continuous data or % for categorical data.

### 3.2 Volumetric analysis

Volumetric measures for the included participants are Figure 1. type 2 diabetes group had significantly lower TIV (p<0.05), and total GM and WM volume (p<0.0001, for both) in comparison to the control group. In contrast, the total CSF volume was significantly higher (p<0.05) in participants with type 2 diabetes than in control participants.

**Figure 1:**
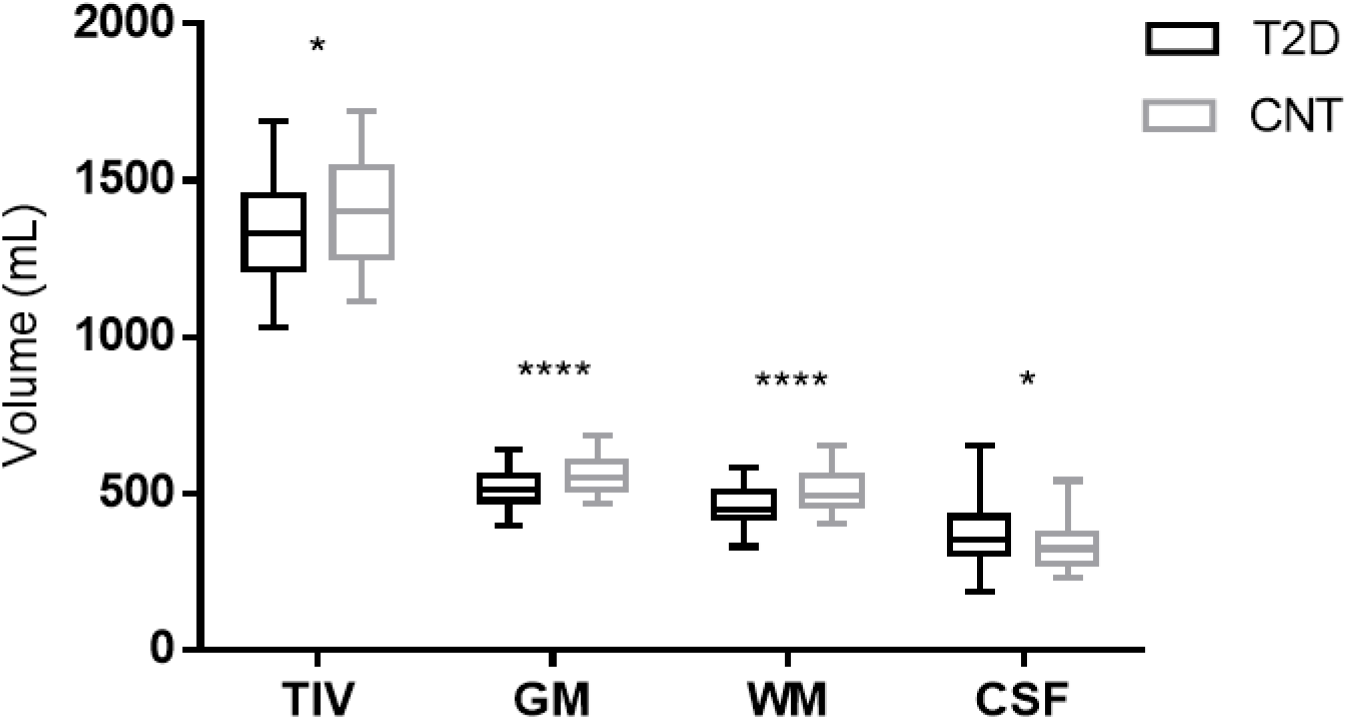
Boxplots of the distribution of total intracranial volume (TIV), total grey matter (GM), white matter (WM) and cerebrospinal fluid (CSF) volumes in each group. A significant difference between groups is indicated by *p<0.05 and ****p<0.0001, assessed with t (TIV, GM and WM) and Mann-Whitney (CSF) tests.

### 3.3 Voxel-based morphometry analysis

VBM analysis of GM showed 18 clusters with significantly decreased GM volume in participants with type 2 diabetes (Figure 2), whereas 5 clusters were located in frontal lobe (D, E, G, L and N), 3 clusters in limbic lobe (B, C and R), 2 clusters in occipital lobe (K and M), 2 clusters in posterior lobe (F and H), 1 cluster in temporal lobe (Q), and 5 clusters were found in sub-lobar regions of insula (A, I, J and P) and of lentiform nucleus (O).

**Figure 2:**
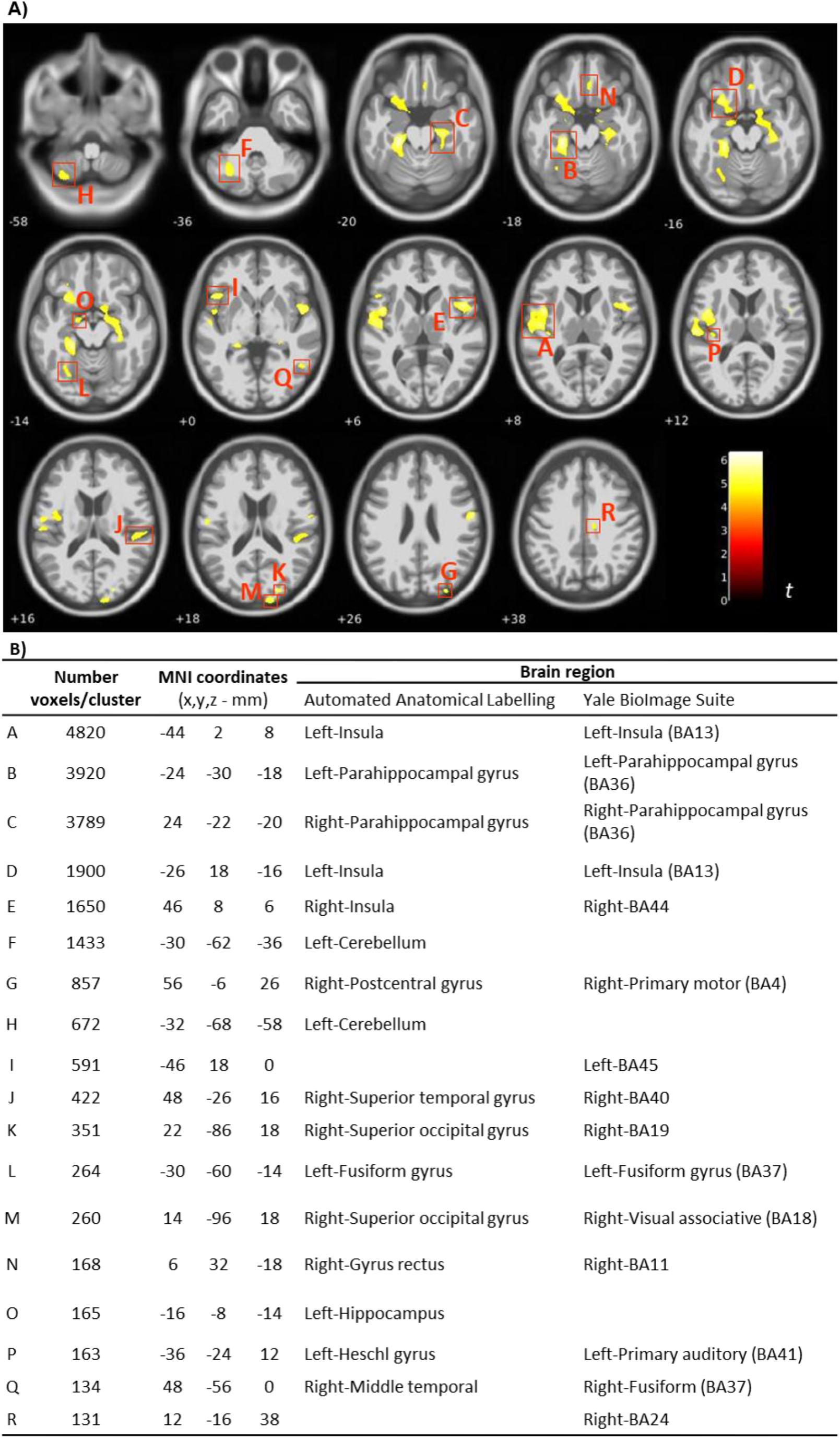
VBM analysis of GM. (A) Statistical map presenting the clusters with significantly decreased GM volume in type 2 diabetes participants, thresholded at p<0.05 with FWE correction for multiple comparisons at voxel level, and with minimum extent cluster size correction (K=114) at cluster level. (B) Labelling of the regions containing clusters with differences, according to their MNI coordinates, based in the Automated Anatomical Labelling and Yale BioImage Suite brain atlases.

### 3.4 Cortical thickness analysis

SBM analysis of cortical thickness revealed 3 clusters with significantly decreased cortical thickness in participants with type 2 diabetes (Figure 3). Clusters were found in temporal lobe (A and C) and in the limbic lobe (B).

**Figure 3:**
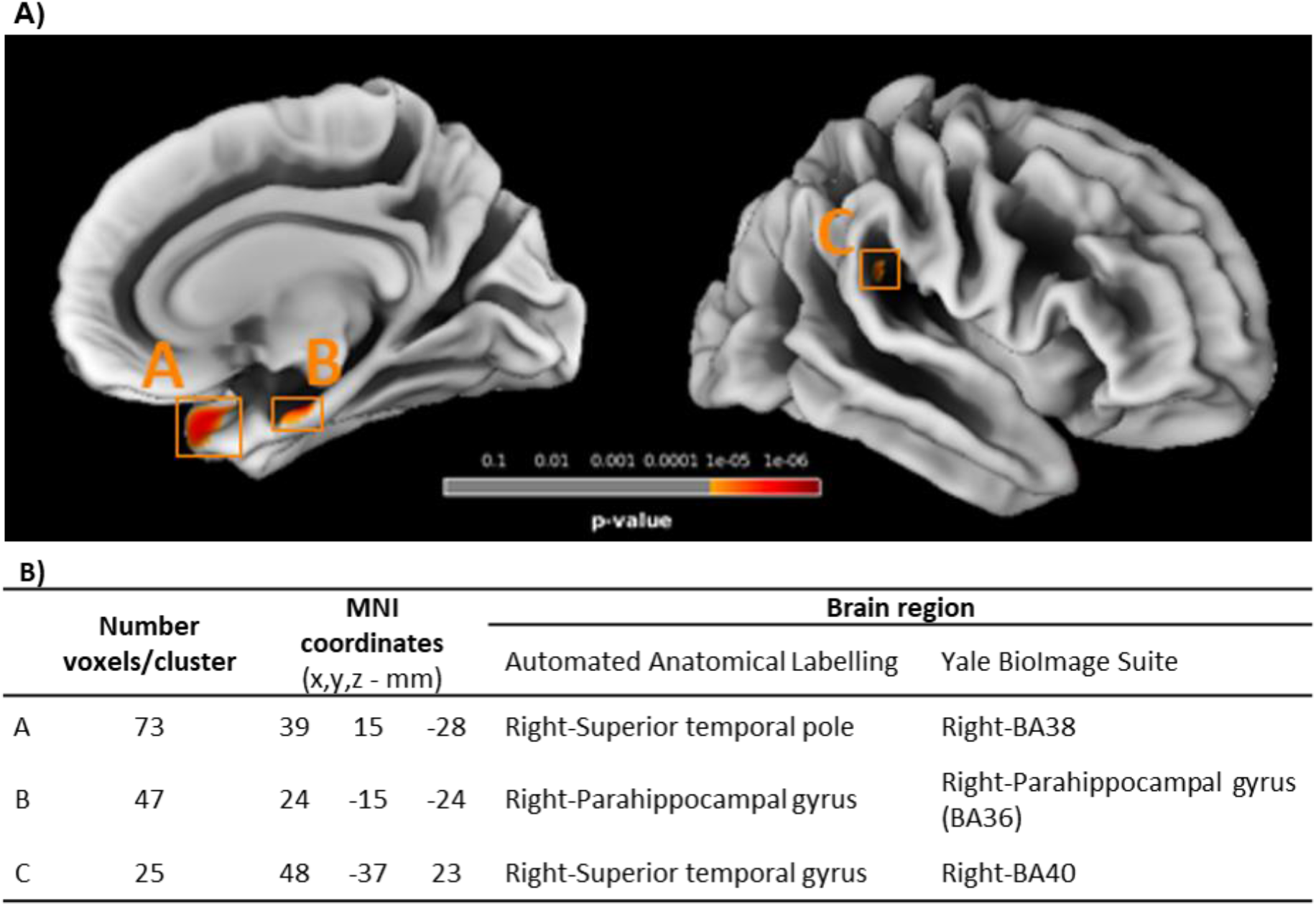
SBM analysis of cortical thickness. (A) Statistical map presenting the clusters with significantly decreased cortical thickness in type 2 diabetes participants, thresholded at p<0.05 with FWE correction for multiple comparisons at vertex level, and with minimum extent cluster size correction (K=25) at cluster level. (B) Labelling of the regions containing clusters with differences, according to their MNI coordinates, based in the Automated Anatomical Labelling and Yale BioImage Suite brain atlases.

**Figure 4:**
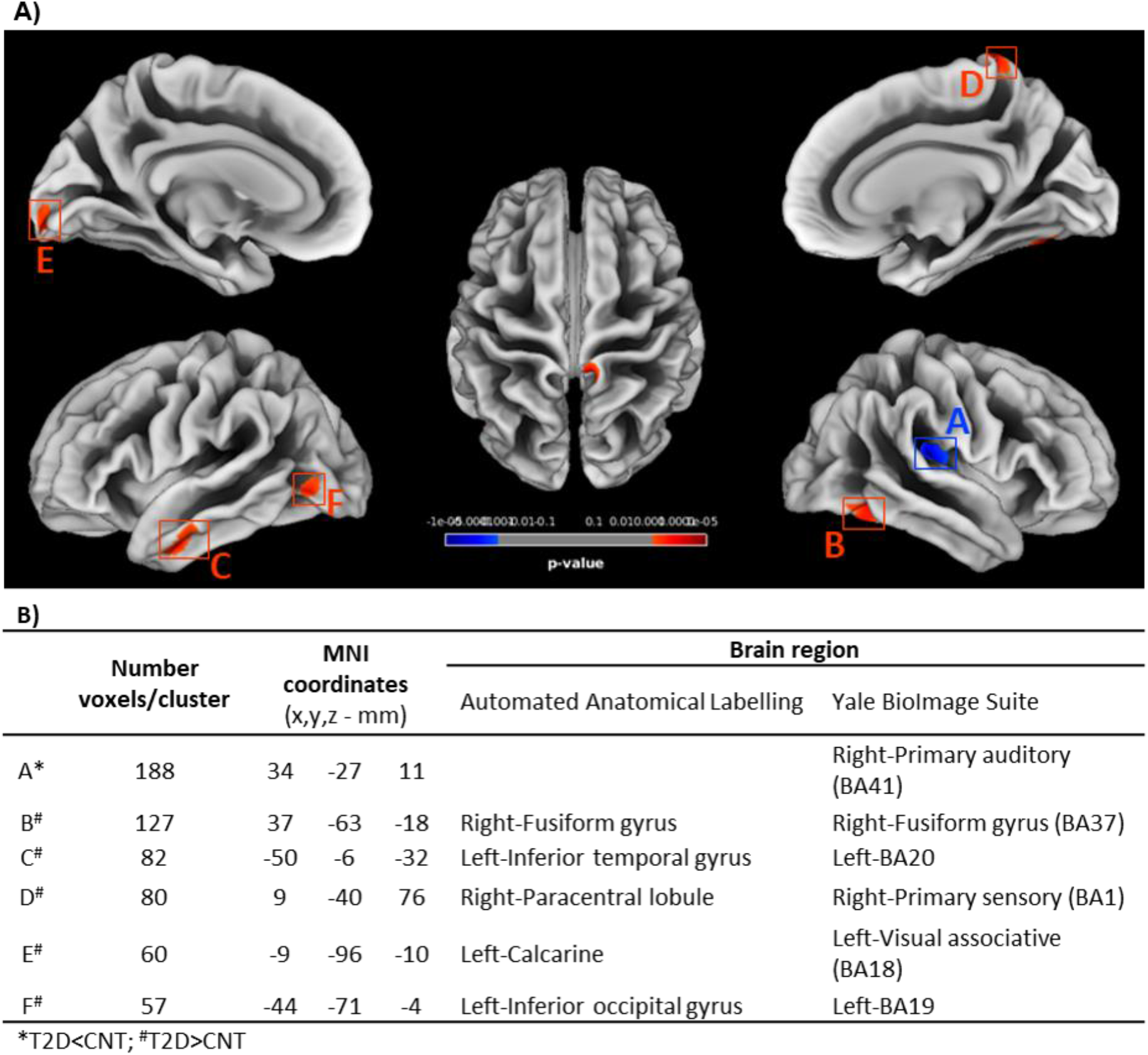
SBM analysis of cortical gyrification. (A) Statistical map presenting the clusters with significantly decreased (*) or increased (#) cortical gyrification index in type 2 diabetes participants, thresholded at p<0.001 at vertex level, and with minimum extent cluster size correction (K=47) at cluster level. (B) Labelling of the regions containing clusters with differences, according to their MNI coordinates, based in the Automated Anatomical Labelling and Yale BioImage Suite brain atlases.

### 3.5 Cortical gyrification analysis

SBM analysis of cortical gyrification index showed a distinctive pattern in particular in sensory regions, revealing 5 clusters with a significant increase in cortical gyrification index in participants with type 2 diabetes and only 1 with lower gyrification index in type 2 diabetes patients (Figure 3). Clusters with increased gyrification index were found in temporal lobes (C and F), posterior lobe (B), parietal lobe (D) and occipital lobe (E). The cluster with decreased gyrification index was in the temporal lobe (A).

The relation between the local gyrification index and clinical data, presented in Table 2, was significant only in clusters with increased gyrification in type 2 diabetes patients. A moderate negative correlation between disease duration and left occipital cortex (cluster F) was found. We also observed positive weak correlations with fasting glucose in left visual cortex (cluster E) and with HbA1C in right fusiform gyrus and left occipital cortex (cluster B and F, respectively).

**Table 2:** Correlations between gyrification index and clinical data

| Clusters |  | A* | B<sup>#</sup> | C<sup>#</sup> | D<sup>#</sup> | E<sup>#</sup> | F<sup>#</sup> |
| --- | --- | --- | --- | --- | --- | --- | --- |
| **Fasting glucose (n=83)** | Spearman rho | -0,198 | 0,12 | 0 | -0,028 | 0,219 | -0,013 |
|  | *p* | 0,07 | 0,275 | 0,999 | 0,799 | 0,044 | 0,908 |
| **Glycated hemoglobin** | Pearson r | -0,06 | 0,232 | 0,051 | 0,059 | 0,103 | 0,266 |
|  | *p* | 0,588 | 0,035 | 0,647 | 0,596 | 0,355 | 0,015 |
| **Age at diagnosis** | Pearson r | -0,027 | 0,095 | 0,189 | 0,138 | -0,042 | 0,012 |
|  | *p* | 0,837 | 0,476 | 0,153 | 0,297 | 0,749 | 0,931 |
| **Disease duration** | Spearman rho | 0,066 | -0,094 | 0,015 | 0,052 | 0,195 | -0,38 |
|  | *p* | 0,617 | 0,479 | 0,91 | 0,698 | 0,139 | 0,003 |

\*T2D<CNT; <sup>#</sup>T2D>CNT

## 4. Discussion

Diabetes is known to impair brain structure and function. Cerebrovascular complications, alterations in glucose homeostasis and insulin signaling, are the factors present in diabetes that most likely affect brain integrity and this is even more critical as neural tissue is highly dependent on glucose to function.

Here we targeted morphometric properties which are not captured by VBM or cortical thickness analyses, by evaluating for the first time cortical gyrification in type 2 diabetes. This feature has been postulated to be more determined by genetic factors mainly during the fetal development [Papini et al., 2020; White et al., 2010; Zilles et al., 1988; Zilles et al., 2013]. Accordingly, the literature has consistently reported altered gyrification index in diseases with genetic or neurodevelopmental basis [Bearden et al., 2009; Gaser et al., 2006; Jou et al., 2010; Palaniyappan and Liddle, 2012]. Nonetheless, the brain nearly triples its size from birth to adulthood and alterations in brain morphology, including in gyrification index, also occur in healthy aging [Hogstrom et al., 2013; Lamballais et al., 2020]. Changes have also been observed in late acquired and diseases with more complex etiology [Lebed et al., 2013; Lin et al., 2007]. These observations raise credit to the notion that changes in gyrification index may occur due to environmental factors [Amunts et al., 1997; Luders et al., 2012].

Regarding the complex etiology of type 2 diabetes, with underlying both genetic and more preponderant environmental factors, we hypothesized that alterations in the brain surface morphology, namely in gyrification, may also occur, in addition to the expected structural changes and functional impairments. To the best of our knowledge, there are no studies assessing the cortical gyrification index in type 2 diabetes in humans.

In this study, we were able to find changes in gyrification index, mainly observed in primary sensory areas. A recent study showing higher correlations between local genetic patterns and cortical folding in somatosensory areas, which suggests evidence that the topology of primary cortex is more constrained by genetics influence than association higher-older cortical areas [Alexander-Bloch et al., 2020]. The fact that the sensory cortex has thicker layers, namely layer IV that receives most projections from the thalamus, might underlie the higher proneness of sensory areas to suffer alterations of gyrification. Ronan et al. have indeed postulated that thalamocortical connectivity is critical to gyrification patterns [Ronan and Fletcher, 2015]. Furthermore, the increase of cortical surface area in sensory regions, as a mechanism to decrease metabolic costs associated with higher degree of local connectivity, might confer additional predisposition to alterations of gyrification in these areas [Alexander-Bloch et al., 2020]. Thus, we believe our finding might be attributed, in part, to a genetic impact, but possibly also to acquired damage to the thalamus, a vulnerable region, and its connections. Importantly, different characteristics of gyrification, such as length and shape of sulci and gyri, might be affected differently by distinct factors. There is evidence of a moderate degree of genetic control over variation in sulcal length, while gyrus-shape features are more susceptible to environmental effects [Atkinson et al., 2015]. This fact further supports the notion that although gyrification is suggested to be more genetically controlled, other factors, namely environmental, might play a role in alterations of gyrification. A study with an animal model of diabetes supports the idea of environmental influence in gyrification. This study showed a severe reduction in cortical convolution in streptozotocin-induced diabetic rats, that showed improvements in this morphologic measure after an antioxidant treatment [Nurdiana et al., 2018]. Therefore, these results leave open the possibility that also the metabolic control might have an effect in gyrification. Notably, in our analysis, we observed significant correlations of the gyrification index with measures reflecting metabolic control. A positive correlation of HbA1c and fasting glucose with increased gyrification in type 2 diabetes suggests a role of hyperglycemia in increasing gyrification index in type 2 diabetes. On the other hand, we observed a negative correlation of disease duration with increased gyrification index in type 2 diabetes. This finding suggests that longer disease duration somehow diminishes the difference of gyrification index in type 2 diabetes relative to healthy controls, which seems to be a summation effect of disease with that of age in gyrification.

In contrast to our results showing decrease in GM volume and thickness, we mainly observed increased gyrification in type 2 diabetes compared to healthy controls. The fact that type 2 diabetes brains showed a higher gyrification index was surprising for us, as we expected it would follow the tendency of other morphometric measures. This suggests a more complex underlying mechanism possibly involved. The diabetic brain is susceptible to a pathological environment including inflammation, oxidative stress, and hypoxia, leads ultimately to axonal damage and cellular death. Brain atrophy is related with neurological insult but gyrification may also be affected by other factors such as connectivity changes [Ronan and Fletcher, 2015]. Interestingly, previous studies showed the same pattern of cortical morphology change, a thinning of the cortex accompanied by an increase in gyrification, in diseases with more genetic or neurodevelopmental basis, such as 22q11.2 deletion [Bearden et al., 2009], Williams syndrome [Gaser et al., 2006], autism [Jou et al., 2010], and schizophrenia [Palaniyappan and Liddle, 2012].

Using more conventional measures we could also show that diabetes impacts brain structure, although literature is not fully consistent, in particular when samples sizes are small. Notably, we could observe highly significant differences in the total volumes of brain tissues, namely the global reduction of GM and WM in type 2 diabetes patients, which is in line with a study with a large cohort of type 2 diabetes patients [Moran et al., 2013]. This reduction was accompanied by a difference also in TIV, which may not be higher due to the increase in CSF values in the type 2 diabetes group. Less studied, increases in CSF volume are related with aging and with dementia [Tanna et al., 1991; White et al., 2010]. Therefore, this increase in our type 2 diabetes population is not surprising. VBM analysis also supported what we found at the whole-brain level, showing only clusters with lower GM volume in type 2 diabetes group. Consistently, we only observed regional cortical thinning in type 2 diabetes group. These results are in accordance with previous studies of brain atrophy using VBM [Moran et al., 2013; Moulton et al., 2015; Wang et al., 2014] and cortical thickness [Brundel et al., 2010; Chen et al., 2015; Chen et al., 2017; Li et al., 2018; Shi et al., 2019] analysis in diabetic populations. In our study, decreased regional GM volumes appeared in several regions in type 2 diabetes patients, mainly in the frontal lobe, as in [Moran et al., 2013; Moulton et al., 2015; Roy et al., 2020], and sub-lobar cortex. Specifically, we observed decreased GM volume in different clusters of the insula, which has been implicated in an overwhelming variety of functions ranging from sensory processing to representation of feelings and emotions [Roy et al., 2020]. This structure was already reported to present impaired neurovascular coupling in an fMRI study with the same cohort [Duarte et al., 2015]. As previously highlighted [Moran et al., 2013; Roy et al., 2020], lower regional GM volume in type 2 diabetes was also observed in our study in the limbic lobe, namely in the cingulate cortex and bilateral parahippocampal gyrus. Furthermore, there was a concomitant GM volume reduction and cortical thinning of the parahippocampal gyrus. A region in the temporal lobe also showed a decrease in cortical thickness [Chen et al., 2017; Li et al., 2018].

## 6. Conclusion

Summing up, we found a novel signature of changed brain structure in type 2 diabetes, the gyrification index, in addition to the expected whole-brain cortical atrophy. Our results regarding the gyrification index are surprising and cannot be simply explained by local neural loss mechanisms. Notably, we found mainly increased gyrification in sensory areas in type 2 diabetes. While a genetic influence should be considered, our correlation results concurrently suggest an influence of metabolic control in alterations of gyrification in type 2 diabetes. Sensory areas are dominated by strong thalamocortical input, and axonal damage might contribute to gyrification changes. This is an important issue to consider in further studies of gyrification in diabetes, addressing the complex etiology of the disease and assessing different underlying factors, including both genetic and environmental determinants. Furthermore, a longitudinal approach will be crucial to investigate in more detail the mechanisms underlying these alterations, ultimately as a potential new biomarker in diabetes management.

## Data Availability

The datasets generated during and/or analysed during the current study are available from the corresponding author on reasonable request.

## Acknowledgments

We thank the participants for their involvement in this study. We also thank C. Ferreira and J. Marques from ICNAS, University of Coimbra, for help with MRI procedures.

## Author contributions

JC, JVD, CM, LG and MCB contributed to conception and design of the study. JVD performed acquisition of data. JC and JVD performed data analysis and interpretation. JC, JVD and MCB wrote the paper. JC, JVD, CM, LG and MCB contributed to the discussion and revised the manuscript.

## Ethics approval statement

The study was approved by the Comissão de Ética da Faculdade de Medicina da Universidade de Coimbra and was conducted in accordance with the declaration of Helsinki. All participants provided written informed consent to participate in the study.

